# Healthcare seeking behavior and delays in case of drug-resistant Tuberculosis patients in Bangladesh: Findings from a cross-sectional survey

**DOI:** 10.1101/2023.04.19.23288805

**Authors:** Md. Zulqarnine Ibne Noman, Shariful Islam, Shaki Akter, Ateeb Ahmad Parray, Dennis G Amando, Jyoti Karki, Zafria Atsna, Dipak Mitra, Shaikh A. Shahed Hossain

**Affiliations:** BRAC James P. Grant School of Public Health (JPGSPH), BRAC university, Dhaka 1212, Bangladesh; EcoHealth Alliance Bangladesh Programs, Institute of Epidemiology, Disease Control and Research (IEDCR), Dhaka 1212, Bangladesh; International Centre for Diarrhoeal Disease Research (icddr,b), Bangladesh, Dhaka, Bangladesh; Department of Public Health, North South University, Dhaka, Bangladesh; Department of International Health, Health systems program, The Johns Hopkins University, Baltimore, USA

## Abstract

**Background:** The emergence of Drug-Resistant Tuberculosis (DR-TB) has become a major threat globally and Bangladesh is no exception. Delays in healthcare seeking, proper diagnosis and initiation of treatment cause continuous transmission of the resistant tubercule bacilli through the communities. This study aimed to assess the different health care-seeking behaviors and delays among DR-TB patients in Bangladesh.

**Method:** A prospective cross-sectional study was conducted from November to December 2018, among 92 culture positive and registered DR-TB patients in four selected hospitals in Bangladesh. Data were collected through face-to-face interviews with survey questionnaire as well as record reviews.

**Result:** Among the 92 study participants, the median patient delay was 7 (IQR 3, 15) days, the median diagnostic delay was 88 (IQR 36.5, 210), the median treatment delay was 7 (IQR 4,12) days, and the median total delay among DR-TB patients was 108.5 (IQR 57.5, 238) days. 81.32% sought initial care from informal healthcare providers. The majority (68.48%) of the informal healthcare providers were drug sellers while 60.87% of patients sought care from more than four healthcare points before being diagnosed with DR-TB. The initial care seeking from multiple providers was associated with diagnostic and total delays.

**Conclusion:** In Bangladesh, DR-TB cases usually seek care from multiple providers, particularly from informal providers, and among them, alarmingly higher healthcare-seeking related delays were noted. Immediate measures should be taken both at the health system levels and, in the community, to curb transmission and reduce the burden of the disease.

## Introduction

Tuberculosis (TB) is a highly transmissible disease caused by *Mycobacterium tuberculosis* bacilli, the second most infectious killer after the COVID-19 pandemic and the 13^th^ leading cause of overall mortality around the world (1). It is estimated that about a quarter of the world’s population is already infected with TB though only 10% of them develop clinical TB during their lifetime (1) (2). The END TB-2030 strategy was developed with a vision of reducing the number of TB death by 90% and reduction of new cases by 80% (3). However, the emergence of drug-resistant tuberculosis (DR-TB) has become a global threat and challenges the End TB-2030 strategy as only one of four new DR-TB cases are now detected while one of two cases got cured (3).

DR-TB is a clinical condition where any of the first-line anti-TB drugs is found resistant. It can be categorized as single-drug-resistant, multi-drug-resistant, or extensive drug-resistant (4). Multi-drug resistant tuberculosis (MDR-TB) is the most common form of DR-TB which is resistant to first-line anti-TB drugs Rifampicin (RFP) and Isoniazid (INR). When one fluoroquinolone and a second-line injectable drug are also resistant, then it is categorized as extensive/extreme drug-resistant tuberculosis (XDR-TB) also known as super-bugs, which may facilitate a global tubercular apocalypse, if not prevented, and intervened timely (5) (6).

About 157 thousand incident cases of DR-TB were notified, and 150 thousand new cases were enrolled in treatment in 2020 according to the Global TB report. However, the estimated number of new cases of DR-TB is 500 thousand each year while only one of the three cases was treated (7). There was a 22% fall in new case detection and a 15% fall in new enrollment in DR-TB treatment from 2019 due to the COVID-19 pandemic. However, only one of three newly developed DR-TB cases each year are enrolled for treatment (1). The undiagnosed, late diagnosed, and lately treated cases cause the continuous spreading of resistant bacilli in the community(8). The high risk of transmission, high case fatality, prolonged treatment, financial, and social burden make DR-TB control strategy more complicated for low and middle socio-income countries like Bangladesh (9). Bangladesh is one of the leading TB burden countries among the top 30 countries (10). In 2020, the laboratory-confirmed number of new DR-TB cases in Bangladesh was 1113, and the incidence rate of DR TB was 2% according to the Global TB report 2020(9).

Early diagnosis of DR-TB and timely initiation of treatment are important. Any delays in the initiation of treatment cause more complications, more disease transmission, and higher mortality. An untreated smear-positive TB case can infect 10 more patients in a year and more than 20 patients during its natural course until death (11). Healthcare-seeking behavior is an influential factor in the diagnosis and prognosis of MDR-TB (12). Patient delays and diagnostic delays depend on patterns of care-seeking of a patient. In South Asia, patients usually visit informal healthcare providers as the first point of care (13). Drug sellers, traditional healers, village doctors, ayurvedic, and homeopaths are recognized as informal healthcare providers in Bangladesh(14). About 60.7% of Bangladeshi patients usually seek first care from pharmacies or traditional providers(15). The number of care-seeking points is usually more as the patients shift from one provider to another until the final diagnosis from the DOTS center which is the diagnostic point of DR-TB. The number of Care seeking points have a greater influence on delay among patients (16). Moreover, among the healthcare providers, the likelihood of initial suspicion about the diagnosis of TB was found low (17). Additionally, social stigma, fear of isolation, and misbelief also cause delays in care-seeking and causes patient delays(18). Gender, education, geography, and socio-economic condition are some contributing factors that determine the care-seeking behavior of DR-TB in sub Saharan Africa(19).

A study in India found a median patient delay of 25 days for uncomplicated pulmonary TB patients (20). In Bangladesh, the median health system delay was found 7.1 weeks and the median treatment initiation delay was found 10 days among MDR-TB patients in a previous study conducted in 2012-2013 (21). WHO recommends no more than 1-day delay for screening to diagnosis and diagnosis to treatment initiation Policy on TB infection control (22). Few authors suggested that a one-month total delay is acceptable while others suggested the total delay should not exceed two months (23). However, very few studies assessed the possible factors related to healthcare-seeking behavior and different delays of DR-TB patients as a whole and particularly from Bangladesh.

The aim of this study was to assess healthcare-seeking behaviors and delays of drug resistance tuberculosis patients. The evidence from this study will be useful for TB control program to take different specific measure at all levels of health systems and in the community level as necessary, to reduce different delays and to implement the END TB strategy by 2030 successfully.

## Methods

### Setting and Study population

We conducted a prospective cross-sectional study, country wide, to reach the objectives. A quantitative survey design was adopted using the prospective cross-sectional approach. In Bangladesh, there are 46 chest clinics, one 250-bedded dedicated TB hospital, several medical college hospitals, and one specialized chest disease hospital for treating TB. We purposefully selected four centers where the flow of TB and DR-TB cases were the most among all centers (24). This study was conducted at the National Institute of Diseases of Chest and Hospital (NIDCH), Dhaka; Mymensingh Tuberculosis and Leprosy hospital; Netrakona Tuberculosis and Leprosy hospital; and Tangail Tuberculosis and Leprosy hospital (Figure-1). A few DR-TB patients (13) were interviewed in nearby Upazilla health complexes (UHC) and adjoining communities of respective catchment areas of enrolled TB hospitals in Netrakona and Tangail also to reach the estimated sample size of 92. The patients from the community were identified from the MDR-TB register of the International Non governmental organization (INGO) - Damien Foundation (DF), as they were under treatment from DF. The NIDCH is the tertiary public hospital in Mohakhali, Dhaka. This is the top referral hospital for DR-TB in Bangladesh. There are 685 beds while 70 beds are reserved for DR-TB patients(25). GF works in 17 countries in Asia. In Bangladesh, they are working in the field of leprosy and TB in collaboration with the National Tuberculosis Control Program (NTP) of Bangladesh. They have three referral hospitals for MDR-TB treatment. This study was conducted in all three referral TB hospitals managed by the DF, which comprises of total 255 beds in Tangail (95 beds), Mymensingh (100 beds), and Netrakona (60 beds) (26).

**Figure 1:**
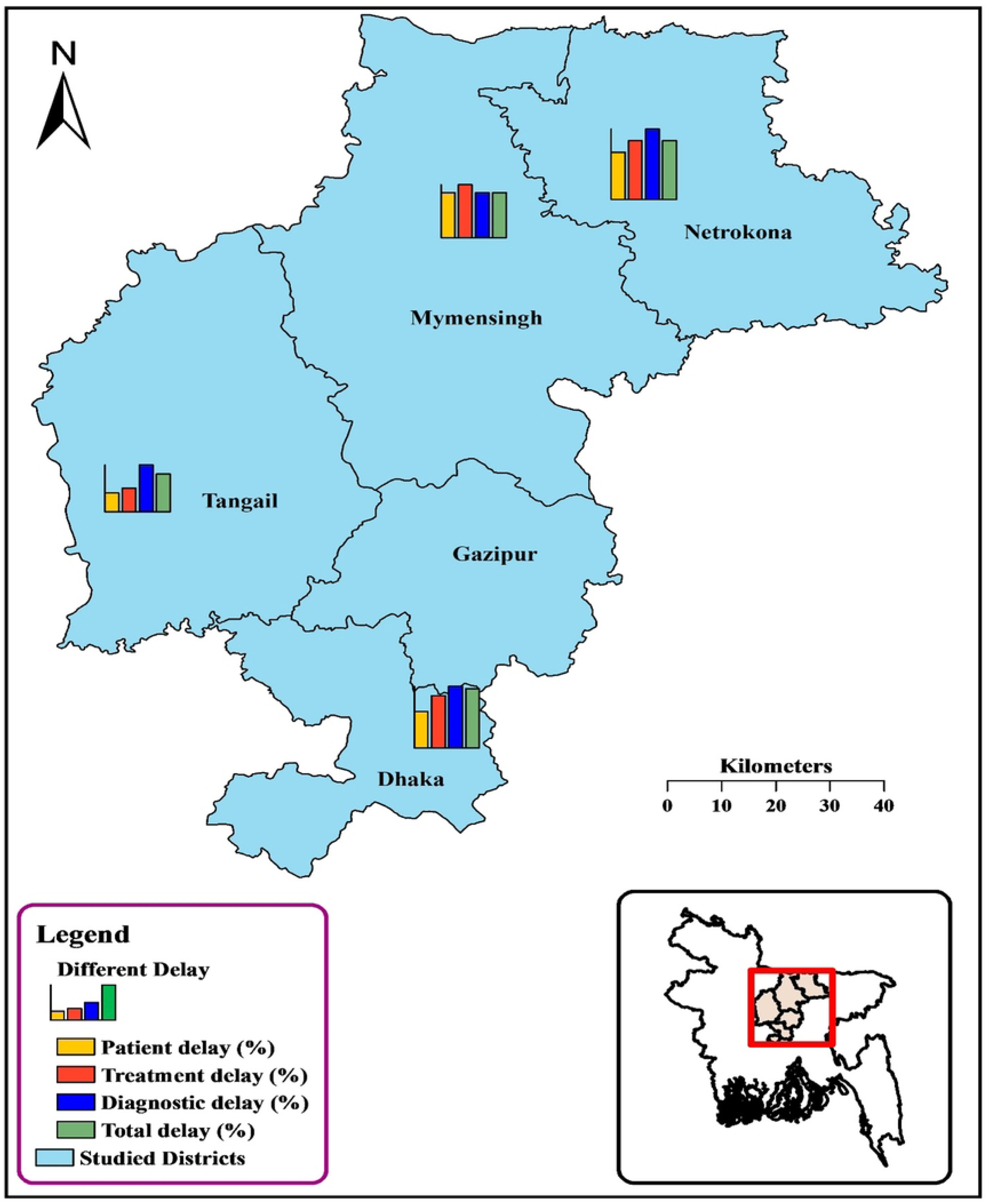
Map of study site locations presenting the distribution of different delays among DR-TB patients in Bangladesh

From 17^th^ November to 7^th^ December 2018, all DR-TB patients admitted to the four selected TB hospitals and community patients available from the near the catchment areas of TB hospitals in Tangail and Netrakona during the time of the survey were considered as the study population. Patients with DR-TB were bacteriologically confirmed, either by GeneXpert or culture. The study comprised patients aged 16 or older who were registered with NTP. All forms of DR-TB treatment registration groups were covered. Patients with severe illnesses were excluded from our study.

A total of 41 patients were from NIDCH, 23 from Mymensingh TB hospital, 9 from Tangail, and 6 from Netrakona TB hospital. Moreover, 13 available DR-TB patients were interviewed from the catchment areas of Netrakona and Tangail TB hospitals.

### Study tools and data collection

A pretested structured survey questionnaire was used to conduct face-to-face interviews. The health care seeking questionnaire was adopted from the WHO document “*Diagnostic and treatment delay in tuberculosis An in-depth analysis of the health-seeking behavior of patients and health system response in seven countries of the Eastern Mediterranean Region*”(27). The questionnaire was translated into the local language Bangla and then again back translated into English to check the accuracy and consistency of the tool. The questionnaire was modified according to the pretesting experiences as per the consensus of the research team. Hospital records were reviewed from the DR-TB card, Sputum register, TB register and Laboratory log to source match the credentials of the hospital admitted study participants as well as participants from catchment areas. Two researchers and three trained research assistants with medical backgrounds who were good in the local language conducted all the interviews. After interviewing each DR-TB patient, the TB-card was reviewed for validation of patient information and for some key information like treatment registration group, date of diagnosis and date of treatment, etc. Any conflict between patient response and data records, record data was preferred to reduce recall bias.

### Operational definitions

Resistance against one or more anti-TB drugs was considered as DR-TB. It can be single-drug resistant, multi-drug resistant, or extensive drug-resistant. Resistance against the first-line anti-TB drug, Rifampicin (RFP) and Isoniazid (INR) was considered as MDR-TB. The patients who had never been treated for TB before were categorized as new MDR-TB. The patients who were previously treated for TB and were declared as cured were considered as relapse MDR-TB. The patients who were previously treated for TB but could not complete or lost to follow up (LTFU) were categorized as re-treatment MDR-TB. When a patient previously received TB treatment but the outcome of the treatment was unknown or undocumented, known as other-MDR-TB (24). Resistant against first-line anti-TB drugs with the resistance of one fluoroquinolone and a second-line injectable drug (aminoglycosides and capreomycin) was categorized as XDR-TB. The participants were asked about the date of the onset of the first symptom regarding TB, the date of the first care-seeking from any providers, the date of diagnosis, and the date of treatment initiation. Patient delay was considered as the time between the onset of symptoms of TB identified by a patient to the first care-seeking from any healthcare provider (20). The time between first care-seeking by a patient and being confirmed diagnosed with DR-TB by GeneXpert/Culture was considered as Diagnostic delay (28). Treatment delay was defined as the time between a patient diagnosed with DR-TB and initiation of DR-TB treatment by healthcare providers(27). The time between a patient first recognizing his/her symptoms of DR-TB and getting initiation of treatment was calculated as total delay (20) (Fig. 2). Delay more than the median value considered as long delay.

**Figure 2:**
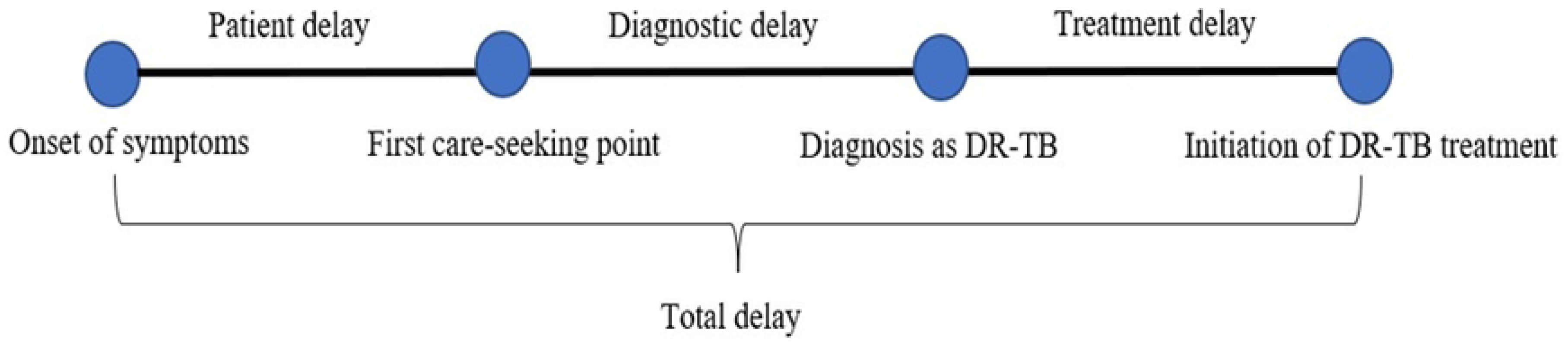
Definition of different delays *(27)* *(28)*.

According to the NTP guideline of Bangladesh, all DR-TB patients are leveled with the different treatment registration groups new cases, relapse cases, failure of treatment by 1^st^ line drug (F1), failure of treatment by 1^st^ line drug due to loss to follow-up, failure of retreatment by 1^st^ line drugs (F2), other cases and XDR-TB. New cases were labeled as those who had no previous history of TB. Relapse cases are those who had a history of TB, but treatment was completed and cured. Other cases were labeled who had a history of TB but no documents of taking or completement of drugs. Doctors, both public and private and DOTS providers are defined as formal healthcare providers in this study. Drug sellers, Village doctors (LMF), Traditional healers, Homeopaths, and Ayurvedic were defined as informal healthcare providers. Self-care at home was also categorized as informal care.

### Data analysis

All data were entered and cleaned through MS excel (Microsoft Office Professional Plus 2021, Microsoft Corporation, Washington, United States) and analyzed through STATA version 16.0 (StataCorp,4905, Lakeway Drive, College Station, Texas 77,845, USA). Descriptive analysis of Socio-demographic characteristics, clinical and personal profile was done using frequency and percentages. Delays were measured using median and range. We performed univariable logistic regression analysis for all predictive factors for different delays and odds ratios (ORs), and 95 % confidence intervals (CI) were calculated. Patient delay was calculated from patient recall. Diagnostic delay was calculated by subtracting patient delay from the date of diagnosis collected from the TB card. Treatment delay was calculated by subtracting the date of diagnosis from the date of treatment initiation collected from the hospital records.

### Ethical approval

Ethical approval was obtained from the institutional review board (IRB) of James P. Grants School of Public Health, BRAC University under reference number **2018-40-lR**. Formal permission was also taken from the head office of Damien Foundation Bangladesh and respective hospital management. Informed written consent was ensured for all respondents over eighteen years after verbal explanation about the study. Informed written consent was taken from parents and assent was taken from respondents under eighteen years of age. Thumbprint was taken, those who were illiterate. The anonymity and confidentiality of all respondents were well-maintained. Personal protective equipment (PPE) including N95 masks was used while interviewing. For community patients, interviews were taken from the nearest directly observed treatment, short-course (DOTS) centers. Transport costs were provided to the community participants.

## Result

Among the total of 92 patients interviewed, 25 (27.17%) were new MDR-TB, 29 (31.52%) were Relapse-MDR-TB, 33 (35.87%) were Re-treatment-MDR-TB, 3 (3.26%) were XDR-TB and 2 (2.17%) were other MDR-TB patients. Among the participants, 69 (75%) were men, 45 (48.91%) patients were 16-34 years of age and most of them 66 (71.74%) were living in rural areas. Most participants were married 76 (82.61%) and engaged in manual work 40 (43.48%). Most of the patients 42 (45.65%) had never attended school. The majority were previously smokers 47 (51.09%) while 15 (16.30%) were found current smokers. The larger portion of the participants 38 (41.30%) had a family income range between 10001-20000 BDT.

Very few of them were alcoholics 6 (6.52%) while 16 (17.39%) had diabetes and 10 (10.87%) had other co-morbid diseases. The history of the previous TB infection was present among 62 (67.39%) patients while 60 (65.22%) had BCG scar proving they were vaccinated.

The most common symptoms presented by the patients at the time of first care-seeking from any healthcare provider were Fever 83 (90.22%) and Cough 78 (84.87%). Other major symptoms were weight loss 61 (66.30%), Haemoptysis 12 (13.04%), Chest pain 23 (25%) and anorexia 10 (10.87%). Three or less symptoms were initially present among most of the 67 (72.83%) patients. The first healthcare provider, they consulted with was an informal provider in most cases 74 (81.32%). The majority of patients 56 (60.87%) sought care from more than four care-seeking points before getting a confirmatory diagnosis (Table-1).

**Table 1:** **Frequency distribution with sociodemographic factors, clinical symptoms, care seeking variables**

| Characteristics | Number of patients (%) |
| --- | --- |
| **Sex** |  |
| Men | 69 (75) |
| Women | 23 (25) |
| **Age (year)** |  |
| 16-34 | 45 (48.91) |
| 35-54 | 23 (25.00) |
| ≥55 | 24 (26.09) |
| **Residence** |  |
| Rural | 66 (71.74) |
| Urban | 22 (23.91) |
| Semi-urban | 04 (4.35) |
| **Marital Status** |  |
| Ever married | 76 (82.61) |
| Never married | 16 (17.39) |
| **Employment** |  |
| Unemployed | 9 (9.78) |
| Sales & service | 16 (17.39) |
| Business | 10 (10.87) |
| Manual work | 40 (43.48) |
| Housewife | 9 (9.78) |
| Other | 8 (8.70) |
| **Educational status** |  |
| Not attended school | 42 (45.65) |
| Primary | 19 (20.65) |
| Secondary & above | 31 (33.70) |
| **Current smoker** |  |
| Yes | 15 (16.30) |
| No | 77 (83.70) |
| **Past smoker** |  |
| Yes | 47 (51.09) |
| No | 45 (48.91) |
| **Family income (BDT)** |  |
| ≤10000 | 32 (34.78) |
| 10001-20000 | 38 (41.30) |
| ≥20000 | 22 (23.91) |
| **Consume alcohol** |  |
| Yes | 6 (6.52) |
| No | 86 (93.48) |
| **DM** |  |
| Yes | 16 (17.39) |
| No | 76 (82.61) |
| **BCG scar** |  |
| Yes | 60 (65.22) |
| No | 21 (22.83) |
| Don't know | 11 (11.96) |
| **History of the previous TB** |  |
| Yes | 62 (67.39) |
| No | 30 (32.61) |
| **Number of care-seeking points** |  |
| ≤5 | 53 (57.61) |
| >5 | 39 (42.39) |
| **Co-morbid disease** |  |
| Yes | 10 (10.87) |
| No | 82 (89.13) |
| **Treatment registration group** |  |
| New, MDR | 25 (27.17) |
| Relapse, MDR | 29 (31.52) |
| Re-treatment, MDR | 33 (35.87) |
| Other, MDR | 2 (2.17) |
| X-DR | 3 (3.26) |
| **Symptoms** |  |
| Fever | 83 (90.22) |
| Cough | 78 (84.78) |
| Weight loss | 61 (66.30) |
| Chest pain | 23 (25.00) |
| Hemoptysis | 12 (13.04) |
| Anorexia | 10 (10.87) |
| Other | 04 (4.35) |
| **Number of initial symptoms** |  |
| ≤3 | 67 (72.83) |
| >3 | 25 (27.17) |
| **Type of first healthcare provider** |  |
| Formal | 17 (18.68) |
| Informal | 74 (81.32) |
| **Number of care-seeking points** |  |
| ≤4 | 36 (39.13) |
| >4 | 56 (60.87) |

The median patient delay was found 7 (IQR 3,15) days. The median diagnostic delay was 88 (IQR 36.5, 210) days and the median treatment delay was 7 (IQR 4,12) days, while the total delay was 108.5 (IQR 57.5, 238) days. Before getting the diagnosis, the median number of care-seeking points was 5 (IQR 4,6) (Figure 2). The mean patient delay was 15.38 days. The mean diagnostic delay was 123.46 days, the mean treatment delay was 9.63 days, while the mean total delay was 148 days.

**Figure 3:**
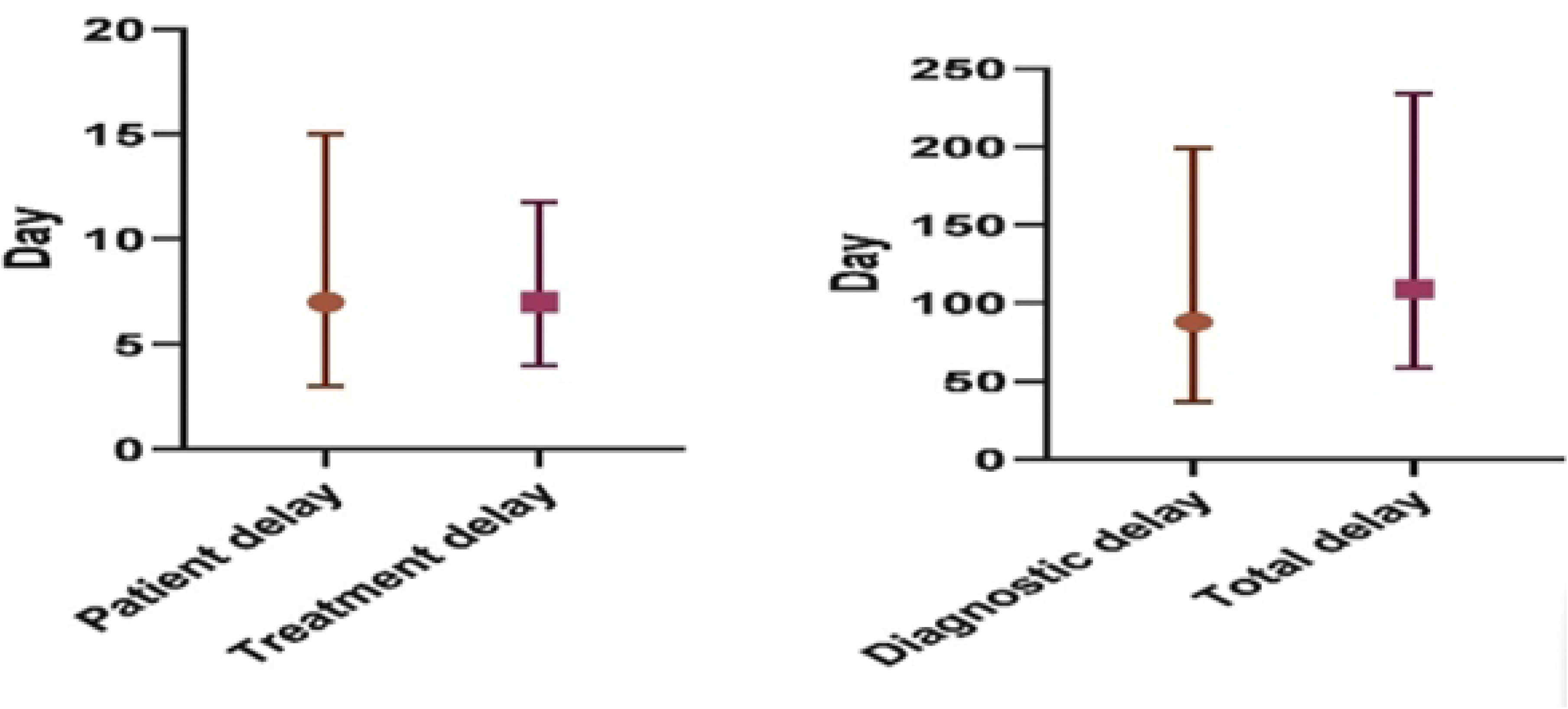
Median and IQR of Different delays among DR-TB patients in Bangladesh

Half of the (54.35%) DR-TB patients had more total delay while patients in NIDCH (63.41%) and Netrakona (62.50%) had the most. Most of the participants from Netrakona had more patients delay (50.00%), diagnostic delay (75.00%) and Treatment delay (62.50%) (Map-1, table-2).

**Table 2:**
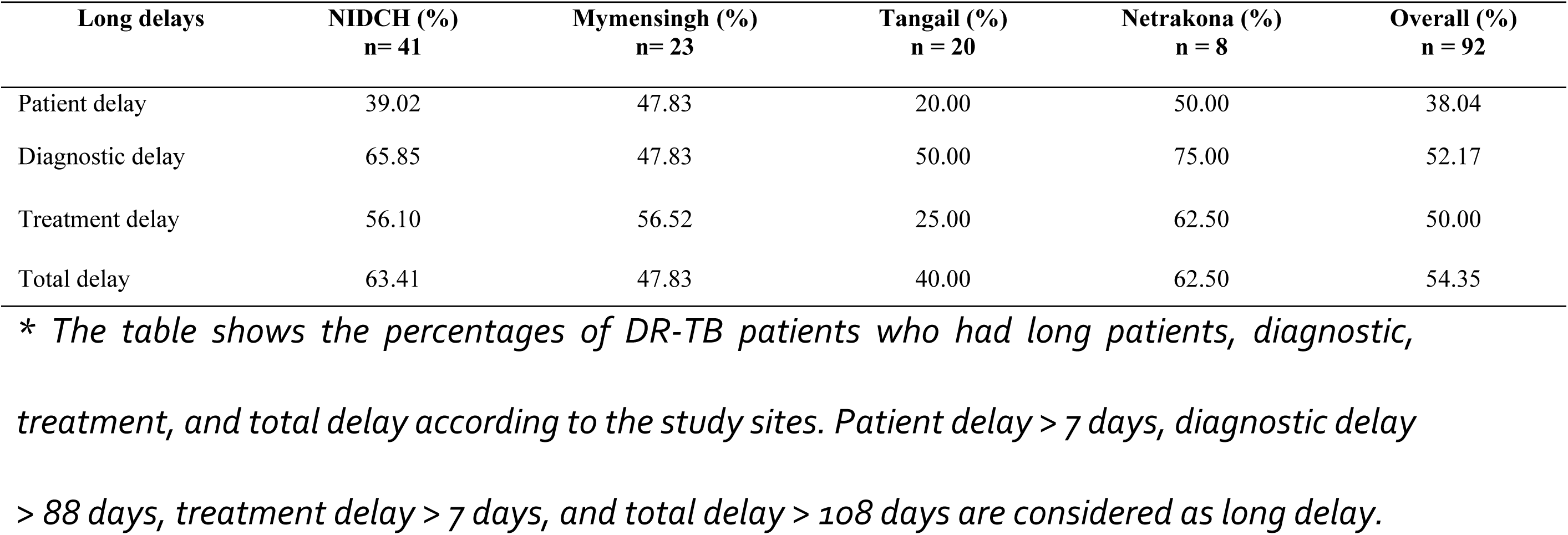
Percentage of DR-TB patients who had long delays according to the study sites*

| Long delays | NIDCH (%)<br>n= 41 | Mymensingh (%)<br>n= 23 | Tangail (%)<br>n = 20 | Netrakona (%)<br>n = 8 | Overall (%)<br>n = 92 |
| --- | --- | --- | --- | --- | --- |
| Patient delay | 39.02 | 47.83 | 20.00 | 50.00 | 38.04 |
| Diagnostic delay | 65.85 | 47.83 | 50.00 | 75.00 | 52.17 |
| Treatment delay | 56.10 | 56.52 | 25.00 | 62.50 | 50.00 |
| Total delay | 63.41 | 47.83 | 40.00 | 62.50 | 54.35 |

\* The table shows the percentages of DR-TB patients who had long patients, diagnostic, treatment, and total delay according to the study sites. Patient delay > 7 days, diagnostic delay > 88 days, treatment delay > 7 days, and total delay > 108 days are considered as long delay.

The median patient delay varied in sex, age, marital status, occupation, monthly family income, and presence of co-morbid diseases. Men had nearly double the median patient delay of 14 (IQR 3,25) days than females 6 (IQR 3,10) days. The young adult group of age (16- 34 years) had more patient delay 7 (IQR 3,15) days than the older age group 35-54 years of age (6.5 days) and ≥55 years of age (5.5 days). The ever-married patients had a more median patient delay of 7 (IQR 3,15) days than the never-married group of 5 (3-14.5) days. Unemployed persons had a median patient delay of 14 (IQR 3,15) days than other professionals such as manual workers, housewives, and businessmen. The person whose occupation was “Sales & Service”, had a less median patient delay of 3.5 (IQR 3,7.5) days. The median patient delay was 4 (IQR 3,7) days among patients who had monthly family income ≥20,000 BDT while those who had monthly family income <20,000 BDT, had a patient delay of 7 (IQR 3,15) days. The patients who had comorbid diseases found less patient delay 4.5 (IQR 3,7) days while others had 7 (IQR 3,15) days (Table 3).

**Table 3:** Socio-demographic characteristics and personal history of DR-TB patients

| Characteristics | N (%) | Patient delay | Diagnosis delay | Treatment initiation delay | Total Delay |
| --- | --- | --- | --- | --- | --- |
| Median delay (IQR) in days* |  |  |  |  |  |
| **Sex** |  |  |  |  |  |
| Women | 69 (75) | 6 (3-10) | 87 (39-224) | 6 (4-9) | 104 (48-241) |
| Men | 23 (25) | 14 (3-25) | 102 (33-180) | 8 (4-14) | 124 (57.5-213.5) |
| **Age (year)** |  |  |  |  |  |
| 16-34 | 45 (48.91) | 7 (3-15) | 101.5 (36-201) | 7 (4-11) | 119 (55-240) |
| 35-54 | 23 (25.00) | 6.5 (3-15) | 96.5 (64-184) | 7 (4-14) | 115 (102-190) |
| ≥55 | 24 (26.09) | 5.5 (1.5-11) | 64.5 (32-221.5) | 8 (5-13.5) | 87 (44-235.5) |
| **Residence** |  |  |  |  |  |
| Rural | 66 (71.74) | 7 (3-14) | 90 (37-226) | 8 (5-13) | 108 (61-250) |
| Urban | 22 (23.91) | 7 (3-30) | 60 (31-129) | 6 (3.5-12) | 68 (45-162) |
| Semi-urban | 04 (4.35) | 3 (1-6) | 140.5(84.5-207.5) | 4.5 (3.5-8) | 149.5 (95-215.5) |
| **Marital Status** |  |  |  |  |  |
| Ever married | 76 (82.61) | 7 (3-15) | 84 (42-206.5) | 7 (4-11) | 106 (62-234.5) |
| Never married | 16 (17.39) | 5 (3-14.5) | 125 (23.5-211.5) | 8 (3.5-13.5) | 141.5 (56-238.5) |
| **Employment** |  |  |  |  |  |
| Unemployed | 9 (9.78) | 14 (3-15) | 52 (37-201) | 6 (4-8) | 61 (55-241) |
| Sales & service | 16 (17.39) | 3.5 (3-7.5) | 80 (24-222) | 8.5 (4-12.5) | 90 (42-236) |
| Business | 10 (10.87) | 7 (3-15) | 64 (30-81) | 7 (4-9) | 91 (47-120) |
| Manual work | 40 (43.48) | 7 (3-10) | 89.5 (60-224) | 8 (4-17) | 104 (78-240) |
| Housewife | 9 (9.78) | 7 (1.5-20) | 106 (62-142) | 6 (5-8) | 134.5 (88.5-189.5) |
| Other | 8 (8.70) | 18 (6-30.5) | 137 (56.5-255) | 4 (3-8) | 208.5 (68-296) |
| **Educational status** |  |  |  |  |  |
| Not attended school | 42 (45.65) | 6 (2-14) | 87 (45-219) | 8 (5-13) | 103 (66.5-241) |
| Primary | 19 (20.65) | 7 (3-15) | 104 (60-222) | 7 (3-13) | 115 (81-162) |
| Secondary & above | 31 (33.70) | 7 (3-15) | 81 (36-180) | 7 (3-11) | 102 (48-241) |
| **Current smoker** |  |  |  |  |  |
| Yes | 15 (16.30) | 7 (4-15) | 125 (60-237) | 7.5 (5-11) | 119 (78-255) |
| No | 77 (83.70) | 7 (3-14.5) | 84 (36-194) | 7 (4-13) | 104 (55-229) |
| **Past smoker** |  |  |  |  |  |
| Yes | 47 (51.09) | 7 (2-10) | 91 (37-231) | 8 (4-14) | 103 (54.5-246) |
| No | 45 (48.91) | 7 (3-15) | 85 (36-180) | 7 (4-10.5) | 111.5 (57.5-213.5) |
| **Family income (BDT)** |  |  |  |  |  |
| ≤10000 | 32 (34.78) | 7 (3-15) | 103 (56-226) | 7 (4-9) | 129 (81-242) |
| 10001-20000 | 38 (41.30) | 7 (3-15) | 60 (19-180) | 8 (3.5-13) | 82 (41-195) |
| ≥20000 | 22 (23.91) | 4 (3-7) | 96 (62-194) | 7 (5-14) | 106.5 (69- |
| **Consume alcohol** |  |  |  |  |  |
| Yes | 6 (6.52) | 3.5 (3-6) | 258.5 (95-297) | 7 (4-7) | 236 (104-303) |
| No | 86 (93.48) | 7 (3-15) | 81 (33-184) | 7.5 (4-13) | 108 (55-229) |
| **DM** |  |  |  |  |  |
| **Yes** | 16 (17.39) | 7 (3.5-20) | 32 (23-64) | 5.5 (3-9) | 63 (33-115) |
| **No** | 76 (82.61) | 7 (3-15) | 98 (60-222) | 8 (4-13) | 112 (78-241) |
| **BCG scar** |  |  |  |  |  |
| **Yes** | 60 (65.22) | 7 (2-10) | 80.5 (37-180) | 7 (3-11) | 102 (58-224) |
| **No** | 21 (22.83) | 7 (3-15) | 97.5 (21-201.5) | 7.5 (5.5-18.5) | 112 (33-229) |
| **Don't know** | 11 (11.96) |  | 180 (60-295) | 7.5 (4-12) | 201.5 (93-291.5) |
| **History of the previous TB** |  |  |  |  |  |
| **Yes** | 62 (67.39) | 7 (3-12) | 105 (62-231) | 8 (4.5-11) | 119 (84-252) |
| **No** | 30 (32.61) | 7 (3-21) | 39 (24-89) | 6 (4-14) | 64.5 (42.5-145) |
| **Number of care-seeking points** |  |  |  |  |  |
| **≤4** | 53 (57.61) | 7 (3-15) | 62 (30-134) | 7 (4-10.5) | 78 (47-150) |
| **>4** | 39 (42.39) | 7 (3-15) | 142 (72-237) | 8 (4-16) | 176 (90-255) |
| **Number of initial symptoms** |  |  |  |  |  |
| **≤3** | 67 (72.83) | 7 (3-15) | 80 (32.5—174.5) | 8 (5-13) | 108 (58-224) |
| **>3** | 25 (27.17) | 6 (3-10) | 143.5(53.5-239.5) | 7 (3.5-8.5) | 150 (55-242) |
| **Type of first healthcare provider** |  |  |  |  |  |
| **Formal** | 17 (18.68) | 8 (3-30) | 39 (23-106) | 5.5 (2.5-10.5) | 67.5 (34-145) |
| **Informal** | 74 (81.32) | 7 (3-14) | 95 (52-226) | 8 (4-13.5) | 109 (63-250) |
| **Co-morbid disease** |  |  |  |  |  |
| **Yes** | 10 (10.87) | 4.5 (3-7) | 67 (33-98) | 6 (4-9) | 75 (55-98) |
| **No** | 82 (89.13) | 7 (3-15) | 90 (37-219) | 7.5 (4-12.5) | 111.5 (57.5-240.5) |
| **Treatment registration group** |  |  |  |  |  |
| **New, MDR** | 25 (27.17) | 7 (3-15) | 36.5 (23-109) | 5 (4-13) | 57.5 (37.5-145) |
| **Relapse, MDR** | 29 (31.52) | 7 (3-10) | 92.5 (30-238.5) | 7.5 (4-11.5) | 102 (47-255) |
| **Re-treatment, MDR** | 33 (35.87) | 5 (3-15) | 105 (70-194) | 8 (5.5-11) | 119 (90-236) |
| **Other, MDR** | 2 (2.17) | 10.5 (7-14) | 119 (37-201) | 6.5 (4-9) | 136 (48-224) |
| **X-DR** | 3 (3.26) | 21 (15-30) | 81 (64-283) | 19.5 (18-21) | 215.5 (115-316) |

The median diagnostic delay was found more among men 102 (IQR 33,180) days than women 87 (IQR 39,224) days. Young adults (18-34 years of age) had more median diagnostic delay of 101.5 (IQR 36, 201) days than other age groups. Urban patients had less median diagnostic delay of 60 (IQR 31, 129) days than rural patients 90 (37, 226) days. Patients who never married found more median diagnostic delay 125 (IQR 23.5, 211.5) days than ever married 84 (IQR 42, 206.5) days. Housewives had more median diagnostic delay 106 (IQR 62,142) days than other professions. Higher educated patients had a less diagnostic delay. Educated up to secondary and higher levels had a median diagnostic delay of 81 (IQR 36, 180) days. Current smokers had more delays 125 (IQR 60, 237) days than non-smokers 84 (IQR 36,194) days. Lower-income patients had a median diagnostic delay of 103 (IQR 56, 226) which was more than higher-income patients. Diabetic patients had less diagnostic delay 32 (IQR 23, 64) days than non-diabetic 98 (IQR 60, 222) days. The patient with a previous TB infection had a median diagnostic delay of 105 (IQR 62, 231) days which was more than those who didn’t have pre-TB 39 (IQR 24, 89) days. The median diagnostic delay of 142 (IQR 72, 237) days was found among those who had number of care-seeking points more than 5 and 62 (IQR 30, 134) days who had the number of care-seeking points ≤5. The patients who sought first care from formal healthcare providers had less diagnostic of delay 39 (IQR 23, 106) days than who got care from informal providers 95 (IQR 51, 226) days. The patients with co-morbid disease had a median diagnostic delay of 67 (IQR 33, 98) days which was less than who didn’t had co morbid diseases 90 (IQR 37, 219) days. The median diagnostic delay of the patients who ha ≤3 initial symptoms was 80 (IQR 32.5, 174.5) days (Table 3).

The median treatment delay was found more among men (8 days), ≥55 years of age (8 days), rural people (8 days), not attend school (8 days), who had previous TB infections (8 days), and who is manual workers (8 days) or occupation was “Sales & Service” (8.5 days). However, the patients who first seek care from formal providers (5.5 days) and patients with co-morbid diseases (6 days) were found to have less median treatment delay (Table 3).

The median total delay was found more among men (124 days), rural patients (108 days), Never married (141.5 days), housewives (134.5 days), manual workers (104 days), family income ≤10,000 BDT (129 days), patients who had pre-TB (119 days), number of care-seeking points more than 5 (176 days) and patients who had initial symptoms more than 3 (150 days). On the other side, the median total delay was comparatively less among ≥55 years of age (87 days), urban patients (68 days), who had diabetes (63 days), who first seek care from formal providers (67.5 days) and who had co-morbid diseases (75 days) (Table 3).

In logistic regression analysis, the patient delay was found significantly associated with never married (OR=4.57, 95 % CI=1.20-17.33), non-diabetic (OR=3.19, 95 % CI=1.01-10.10) (Table-4). The diagnostic delay was significantly associated with the number of care-seeking points more than 4 (OR=3.33, 95% CI= 1.38-8.03), non-diabetic (OR=10.16, 95% CI= 2.15-47.90). No variable was found significantly associated with treatment delay. However, total delay was significantly associated with being non-diabetic (OR=5.65, CI= 1.49-21.45), and the number of care-seeking points was more than 4 (OR=3.79, 95% CI= 1.55-9.24).

**Table 4:**
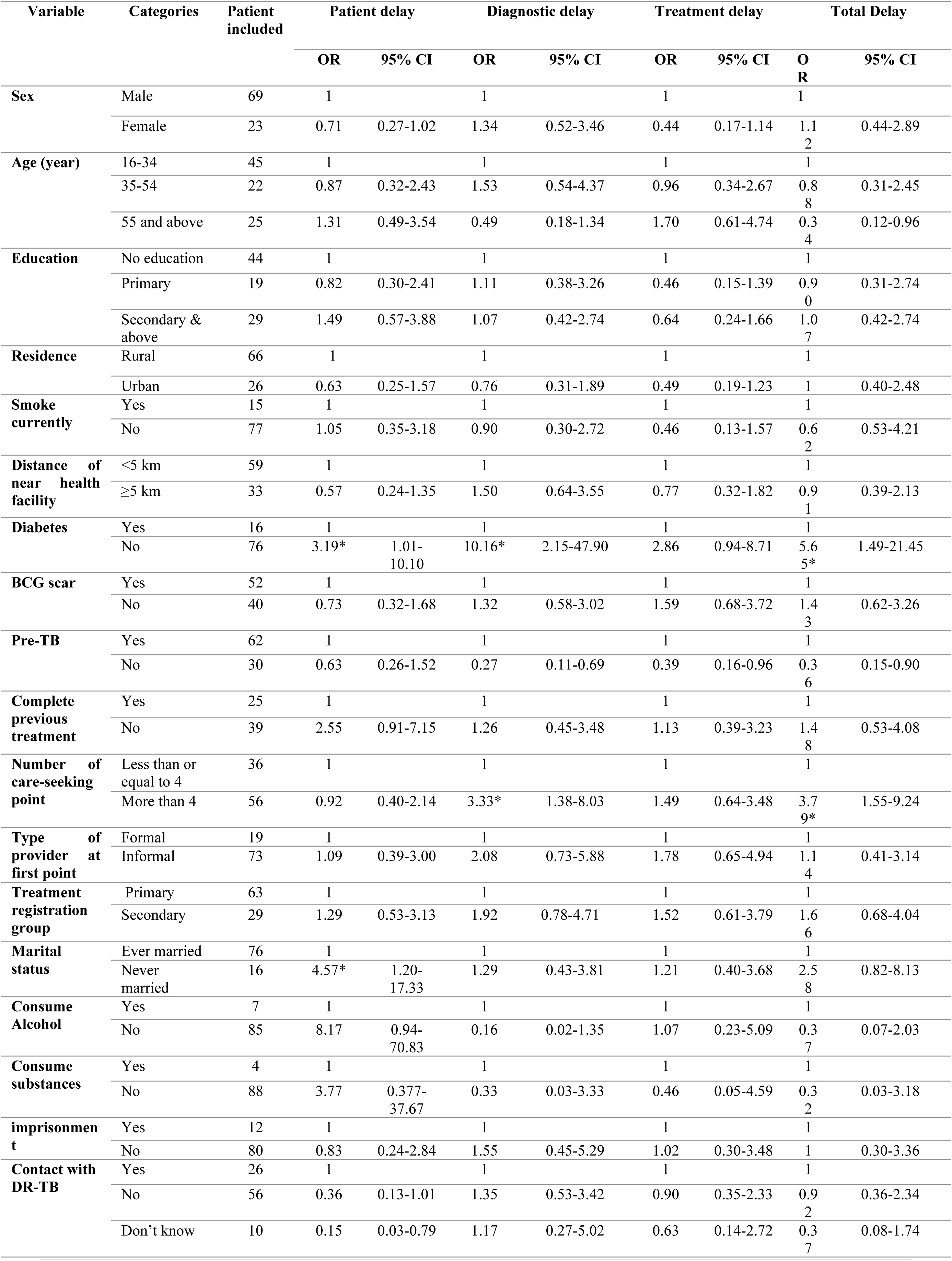
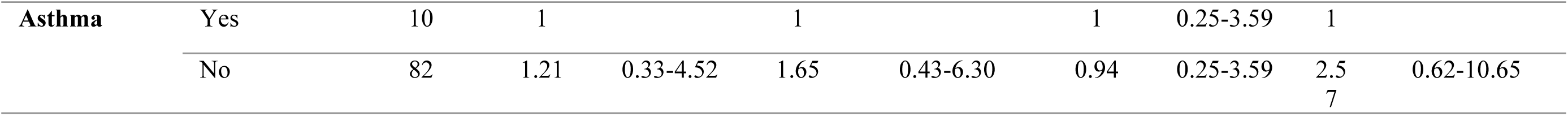
Logistic regression analysis of the determinants of the patient, diagnostic, treatment, and total delay among the DR-TB patients

| Variable | Categories | Patient included | Patient delay |  | Diagnostic delay |  | Treatment delay |  | Total Delay |  |
| --- | --- | --- | --- | --- | --- | --- | --- | --- | --- | --- |
|  |  |  | OR | 95% CI | OR | 95% CI | OR | 95% CI | OR | 95% CI |
| Sex | Male | 69 | 1 |  | 1 |  | 1 |  | 1 |  |
|  | Female | 23 | 0.71 | 0.27-1.02 | 1.34 | 0.52-3.46 | 0.44 | 0.17-1.14 | 1.12 | 0.44-2.89 |
| Age (year) | 16-34 | 45 | 1 |  | 1 |  | 1 |  | 1 |  |
|  | 35-54 | 22 | 0.87 | 0.32-2.43 | 1.53 | 0.54-4.37 | 0.96 | 0.34-2.67 | 0.88 | 0.31-2.45 |
|  | 55 and above | 25 | 1.31 | 0.49-3.54 | 0.49 | 0.18-1.34 | 1.70 | 0.61-4.74 | 0.34 | 0.12-0.96 |
| Education | No education | 44 | 1 |  | 1 |  | 1 |  | 1 |  |
|  | Primary | 19 | 0.82 | 0.30-2.41 | 1.11 | 0.38-3.26 | 0.46 | 0.15-1.39 | 0.90 | 0.31-2.74 |
|  | Secondary & above | 29 | 1.49 | 0.57-3.88 | 1.07 | 0.42-2.74 | 0.64 | 0.24-1.66 | 1.07 | 0.42-2.74 |
| Residence | Rural | 66 | 1 |  | 1 |  | 1 |  | 1 |  |
|  | Urban | 26 | 0.63 | 0.25-1.57 | 0.76 | 0.31-1.89 | 0.49 | 0.19-1.23 | 1 | 0.40-2.48 |
| Smoke currently | Yes | 15 | 1 |  | 1 |  | 1 |  | 1 |  |
|  | No | 77 | 1.05 | 0.35-3.18 | 0.90 | 0.30-2.72 | 0.46 | 0.13-1.57 | 0.62 | 0.53-4.21 |
| Distance of near health facility | <5 km | 59 | 1 |  | 1 |  | 1 |  | 1 |  |
|  | ≥5 km | 33 | 0.57 | 0.24-1.35 | 1.50 | 0.64-3.55 | 0.77 | 0.32-1.82 | 0.91 | 0.39-2.13 |
| Diabetes | Yes | 16 | 1 |  | 1 |  | 1 |  | 1 |  |
|  | No | 76 | 3.19* | 1.01-10.10 | 10.16* | 2.15-47.90 | 2.86 | 0.94-8.71 | 5.65* | 1.49-21.45 |
| BCG scar | Yes | 52 | 1 |  | 1 |  | 1 |  | 1 |  |
|  | No | 40 | 0.73 | 0.32-1.68 | 1.32 | 0.58-3.02 | 1.59 | 0.68-3.72 | 1.43 | 0.62-3.26 |
| Pre-TB | Yes | 62 | 1 |  | 1 |  | 1 |  | 1 |  |
|  | No | 30 | 0.63 | 0.26-1.52 | 0.27 | 0.11-0.69 | 0.39 | 0.16-0.96 | 0.36 | 0.15-0.90 |
| Complete previous treatment | Yes | 25 | 1 |  | 1 |  | 1 |  | 1 |  |
|  | No | 39 | 2.55 | 0.91-7.15 | 1.26 | 0.45-3.48 | 1.13 | 0.39-3.23 | 1.48 | 0.53-4.08 |
| Number of care-seeking point | Less than or equal to 4 | 36 | 1 |  | 1 |  | 1 |  | 1 |  |
|  | More than 4 | 56 | 0.92 | 0.40-2.14 | 3.33* | 1.38-8.03 | 1.49 | 0.64-3.48 | 3.79* | 1.55-9.24 |
| Type of provider at first point | Formal | 19 | 1 |  | 1 |  | 1 |  | 1 |  |
|  | Informal | 73 | 1.09 | 0.39-3.00 | 2.08 | 0.73-5.88 | 1.78 | 0.65-4.94 | 1.14 | 0.41-3.14 |
| Treatment registration group | Primary | 63 | 1 |  | 1 |  | 1 |  | 1 |  |
|  | Secondary | 29 | 1.29 | 0.53-3.13 | 1.92 | 0.78-4.71 | 1.52 | 0.61-3.79 | 1.66 | 0.68-4.04 |
| Marital status | Ever married | 76 | 1 |  | 1 |  | 1 |  | 1 |  |
|  | Never married | 16 | 4.57* | 1.20-17.33 | 1.29 | 0.43-3.81 | 1.21 | 0.40-3.68 | 2.58 | 0.82-8.13 |
| Consume Alcohol | Yes | 7 | 1 |  | 1 |  | 1 |  | 1 |  |
|  | No | 85 | 8.17 | 0.94-70.83 | 0.16 | 0.02-1.35 | 1.07 | 0.23-5.09 | 0.37 | 0.07-2.03 |
| Consume substances | Yes | 4 | 1 |  | 1 |  | 1 |  | 1 |  |
|  | No | 88 | 3.77 | 0.377-37.67 | 0.33 | 0.03-3.33 | 0.46 | 0.05-4.59 | 0.32 | 0.03-3.18 |
| imprisonment | Yes | 12 | 1 |  | 1 |  | 1 |  | 1 |  |
|  | No | 80 | 0.83 | 0.24-2.84 | 1.55 | 0.45-5.29 | 1.02 | 0.30-3.48 | 1 | 0.30-3.36 |
| Contact with DR-TB | Yes | 26 | 1 |  | 1 |  | 1 |  | 1 |  |
|  | No | 56 | 0.36 | 0.13-1.01 | 1.35 | 0.53-3.42 | 0.90 | 0.35-2.33 | 0.92 | 0.36-2.34 |
|  | Don't know | 10 | 0.15 | 0.03-0.79 | 1.17 | 0.27-5.02 | 0.63 | 0.14-2.72 | 0.37 | 0.08-1.74 |
| **Asthma** | Yes | 10 | 1 |  | 1 |  | 1 | 0.25-3.59 | 1 |  |
|  | No | 82 | 1.21 | 0.33-4.52 | 1.65 | 0.43-6.30 | 0.94 | 0.25-3.59 | 2.57 | 0.62-10.65 |

## Discussion

Healthcare-seeking patterns determine the progression, prognosis, and outcome of DR-TB patients. Choice of healthcare has a key role in the journey to get a cure. The first care seeking point is important as well as the number of providers. Bangladesh has a pluralistic health system where the coexistence of different stakeholders, including formal and informal healthcare providers, is usual(29). The referral system is not well-established, especially between informal to formal providers. This study revealed that most of the DR-TB patients (81.32%) were initially visited by informal providers, dominantly drug sellers (68.48%). Other informal providers were village doctors, traditional healers, homeopaths, ayurvedic, etc. Similar findings were found in other studies regarding care-seeking behaviors of TB/MDR-TB patients in Bangladesh(14)(30). A study among TB patients in India, Zimbabwe, and Ethiopia found that two-thirds of them sought first care from non-qualified professionals(31)(32)(33). Informal care providers usually treat with medicine without suggesting any investigations(15). Prescription of antibiotics irrationally is also common among them(34). Also, they don’t refer patients to formal providers or DOTS centers. So, a large portion of the patients had more diagnostic delays found by this study. Patients seeking initial care from informal providers were found to have more than twice the median diagnostic delay, as those who seek care from formal providers. A previous study among TB patients found more health system delays (52%) who seek care from informal providers than formal providers (16%)(34). Policymakers should keep in mind this larger portion of the cake informal caregivers for effective control of DR-TB. Awareness programs should introduce, train them, and establish a referral system between informal to formal caregivers to reduce delays.

The study calculated the number of care-seeking points from the initial symptom of DR-TB to confirmatory diagnosis. The median number of care-seeking points for DR-TB patients was found 5 which is more than in other countries like the USA-4, India-2, and Zimbabwe-3 (35) (33) (32). Care seekers from informal providers had more care-seeking points before diagnosis. More care-seeking points cause more diagnostic and health system delays. Ultimately, these phenomena cause a longer period of DR-TB transmission in the communities. Preventing this vicious cycle is of utmost need to control DR-TB in low resource countries like Bangladesh.

The median time between the onset of the first symptom of DR-TB and the first care-seeking was found 7 days in this study, which is the patient delay. No other study in Bangladesh calculated the patient delays among DR-TB patients. However, studies from other countries found more median patient delays among DR-TB/uncomplicated TB patients such as Zimbabwe 26 days, Ethiopia 35 days, China 58 days (95% of cases), USA- 25 days, and India 15 days(33)(31)(6)(36)(37). One of the reasons for the low patient delay found in our study is the operational definition. This study defined ‘patient delay’ as the time between the appearance of the first symptom to the first consultation with healthcare providers irrespective of their types formal or informal, whether most of the study considered only formal providers(38). The healthcare system of many countries is not as pluralistic as Bangladesh. Seeking care from drug sellers is common in Bangladesh as there is no strict monitoring system though there is a provision of not selling drugs without the prescription of qualified professionals. Two out of five people in Dhaka the capital city of Bangladesh seek care from community pharmacies(39). From the patient perspective, at the community level, pharmacies/informal providers are nearer, less time-consuming, and more cost-effective than formal healthcare settings.

We found a median diagnostic delay of 88 days among the study participants. A previous study (2012-13) in Bangladesh among MDR-TB patients found a health system delay of 7.1 weeks while the provider’s delay was only 4 weeks which is much lesser than the findings of this study(10). There was a possible role of informal providers for this variation. Very few participants (10.8%) in that study sought initial care from informal providers compared to the findings of our study (81.32%). Female participants (75% vs 33.3%), Rural participants (71.74% vs 49.3%), and illiterates (45.65% vs 22%) were more in our study than Rifat et al. Moreover, 27.17% of the participants of this study were categorized as new MDR-TB while the previous study had only 2.4 %(40). The median diagnostic delay among MDR-TB patients in China was 84 days while in Zimbabwe 97 days(41)(6)(33) which was very similar. A previous study among TB patients found a diagnostic delay of 68.5 days in Bangladesh which is less than our study(15). Seeking care from informal providers at the first point, seeking care from multiple healthcare providers, or the tendency to switch providers several times contributes to diagnostic delay. On top of that, qualified healthcare providers occasionally didn’t suggest investigations necessary for diagnosing DR-TB(11). The diagnostic delay is a key indicator of the health system. Reducing diagnostic delays through strengthening a pluralistic health system is a challenge for Bangladesh.

This study found a median treatment initiation delay of 7 days. The median treatment initiation delays found previous studies was 9 days in China(42), 8 days in Gujrat, India(43) 13 days in Myanmar(44), 10 days in Bangladesh(21), 10-22 days in South Africa(45), 9-17 days in Russia(46), 7 days in Bhopal, India(47). The treatment initiation delays mostly occur due to repetition of confirmatory tests, baseline investigations before starting treatment, and sometimes an inadequate number of resident beds for DR-TB in tertiary hospitals(21). Also, the patient’s seriousness about the disease due to lack of awareness and motivation is another possible cause of delay(48). Treatment delay results in poor prognosis and risk of transmission of DR-TB bacilli to the community. However, the treatment initiation delays can be reduced by ensuring an adequate number of beds for DR-TB in all tertiary hospitals, ensuring proper counseling after diagnosis, and suggesting baseline investigations after admission of DR-TB patients into hospitals.

The median total delay was 108.5 days in this study. Previous studies found the median diagnostic delay of 57 days in the USA(36), 120 days in China, 100 days in Pakistan(27), and 132 days in Zimbabwe. Rifat et al found a median of 7.1 weeks of health system delay among MDR-TB patients in Bangladesh in 2015(21). Another study among TB patients in Bangladesh found a total delay of 12 weeks(34). The patient delays, diagnostic delays, and treatment delays contribute to the length of total delays. Both patients and health systems are responsible for long total delays. During these delays, resistant tuberculosis bacilli are continuously spread from person to person. The patient’s lung condition can worsen during this period. Thus, the prognosis of the disease becomes poor which contributes to treatment failure, higher mortality, and economic loss. The chance of fatality is very high in case of a long total delay(21).

Being non-diabetic was significantly associated with patient, diagnostic, and total delays. In Bangladesh, most (over 50%) diabetic patients are registered with Bangladesh Diabetic Somity (BADAS) and seek care (95.2%) from private facilities of BADAS while only 4.3% seek care from informal providers(49). This scenario can clarify why diabetic patients had fewer delays than non-diabetics in Bangladesh. The patient’s delay was significantly associated with never being married in this study. Unmarried people may have a lack of self-motivation and social support, this may be the possible reason for more patient delays among them(50). The number of care-seeking points more than 4 was found significantly associated with diagnostic and total delays. This is obvious that more care-seeking points will contribute to more delays.

The strength of the study is that the investigators interviewed patients face to face. Thus, the information regarding different delays could easily validate with the patient’s memory and record files. DR-TB patients are not available in abundance in the hospitals due to the low case detection rate in Bangladesh(51)(52). The study covered both rural and urban populations as well as non-government and government TB hospitals within certain geographic regions. There is also a possibility of recall bias regarding the patient delays and healthcare-seeking behavior, as researchers needed to rely on the patient’s memory. However, most of the respondents were drawn from the intensive phase of treatment, so the recall period was lesser than community patients. Sometimes few medical records were found missing thus it was difficult to identify the date of diagnosis or treatment initiation. However, researchers had made an effort to minimize the discrepancies by checking all medical records available to patients and service records of hospitals. Few data regarding delays were missing as some patients could not remember the date of their first care seeking. So, calculation of total delays also was not possible for those patients. This missing data on patient delay did not affect the overall outcome of the study.

## Conclusion

Most of the DR-TB patients sought initial healthcare from informal providers while most of the patients seek care from more than five care-seeking points leading to more patient and diagnostic delays. Patient, diagnostic, and treatment initiation delays among the patients were more than recommended resulting in the transmission of the drug-resistant bacilli in the community and increasing the burden of the disease. Community-based interventions are necessary to ensure needful healthcare from formal providers and to reduce the number of care-seeking points before diagnosis. Appropriate training and awareness among healthcare providers could reduce diagnostic delays. This study also recommends ensuring the availability of the Rapid diagnostic test e.g., GeneXpert in primary health care centers. The evidence could help the policymakers, public health specialists, and healthcare providers to improve the TB control program, ensure mass awareness to reduce patient delay, strengthen the health system to reduce diagnostic delay, and ensure early admission to reduce the treatment delay with a vision to the successful implementation of the “END TB 2030” strategy.

## Data Availability

The data are unavailable to the public because they contain personal identifying information and very sensitive information about drug-resistant Tuberculosis patients in Bangladesh who are subjected to extreme stigma and discrimination. Ethical approval for the conduct of the study required that all data, including the locations of patients, be de-identified, imposed by Institutional Review Board (IRB) of BRAC James P Grant School of Public Health. For questions related to data availability please reach out to the principal Investigator - Dr. Shaikh A. Shahed Hossain.

## Acknowledgements

The study was conducted under Summative Learning Project (SLP), as a partial requirement of MPH degree at BRAC James P Grant School of Public Health, BRAC University. This work has been funded by collectively by BRAC James P Grant School of Public Health, BRAC University and TDR under grant number: B40297, the Special Programme for Research and Training in Tropical Diseases, which is hosted at the World Health Organization and co sponsored by UNICEF, UNDP, the World Bank and WHO. We are also indebted towards Dr. Aung, Country director, Damien foundation and Dr. Abdul Hamid Selim, Sr. Scientist, National Tuberculosis and Leprosy control Programme, Ministry of Health and family welfare, Govt. of People’s Republic of Bangladesh for their full-on cooperation during the timeframe of the study.

